# AI-VDT can Help in Detecting Primary Lung Cancer

**DOI:** 10.1101/2022.04.26.22274299

**Authors:** Kazuhiro Suzuki, Yujiro Otsuka, Kota Imashimizu, Kazunori Hata, Kenji Suzuki

## Abstract

The current study shows that measuring volume doubling time could accelerate the detection of primary lung cancer in chest CT imaging. Thirty tumors were selected from surgical cases of primary lung cancer at a university hospital in Japan, and the CT scan data and radiology reports were extracted retrospectively. The CT scan data were processed by a commercial pulmonary nodule AI and volume doubling time was calculated for each historical study time point. 43% of the 30 tumors had an VDT below 400 days in earlier study than the tumor was reported as a nodule on the radiology report. The average days of earlier detection was 299 days. Interpolation of the detailed mortality reduction data from the NELSON study predicted a 5.6% reduction in 10-year mortality from lung cancer.

## Introduction

Previous studies on the clinical usefulness of artificial intelligence (AI)-based pulmonary nodule display programs (“pulmonary nodule AI”) have mainly focused on their effectiveness in improving detection rates in the context of assisting diagnostic imaging, and more specifically, on their effectiveness in preventing missed findings by diagnostic imaging physicians. Therefore, ground truth, the denominator of the detection rate, is the nodule that is judged by a skilled radiologist to be a nodule to be detected for diagnostic imaging purposes, and is often indeterminate with respect to its malignancy. While this is a goal-oriented approach for the purpose of measuring the effectiveness of diagnostic imaging support, it can only be indirectly inferred as a benefit from the reduction of missed cases when discussing patient benefit.

In this regard, evidences (NLST, COSMOS, NELSON, etc.) that improved sensitivity in lung cancer screening reduces the mortality rate of lung cancer patients had made it clear that the prevention of missed diagnostic imaging results in patient benefit. For example, in the National Lung Screening Trial (NLST) in the U.S., screening with computed tomography (CT) three times a year in 53,454 individuals at high risk for lung cancer reduced the median mortality rate by 6.5% compared to screening with chest radiography. years, and more recently, mortality from lung cancer at a median of 5.5 and 6.0 years was found to be up to 19% lower with CT screening than with chest radiography screening [1,2]. In addition, the Dutch-Belgian lung-cancer screening The Dutch-Belgian lung-cancer screening trial (Nederlands-Leuvens Longkanker Screenings A follow-up study of Onderzoek [NELSON]) showed that lung cancer screening with CT scan reduced mortality from lung cancer by 26% over 10 years [3].

Recent advances in MDCT (Multi Detector-row CT) had enabled early detection of small lung cancer lesions and early treatment. However, the amount of image data obtained by MDCT was enormous (several hundred images or more per patient), making it difficult to utilize the data beyond the visual diagnosis and measurement in a two-dimensional plane, which were commonly used in clinical practice. On the other hand, artificial intelligence can process all voxel data without subjectivity or bias, and can calculate parameters in four or more dimensions that are considered clinically useful, such as tumor volume measurement, in addition to tumor detection. In other words, it is still possible to find effectiveness beyond the clinical significance of conventional diagnostic imaging by utilizing the characteristics of AI, which is beyond the capability of most diagnostic imaging physicians to capture medical images as information and store lesions as voxel-by-voxel electronic information.

For example, while the measurement of tumor volume was recognized for its clinical usefulness, in actual clinical practice, it had limitations that were extremely difficult to improve in terms of accuracy and reproducibility of human manual segmentation. Accuracy and reproducibility were both critically important in diagnostic imaging, and the current situation in Japan was that not all cases were measured to determine tumor volume for the sake of operational efficiency.

The volume doubling time (VDT) is an index for evaluating malignancy using tumor volume. Where tumor malignancy is measured in terms of growth rate, invasiveness, and metastatic potential, VDT is a direct indicator of malignancy, and its usefulness had been demonstrated [4,5]. The Fleischner Society, in its lung nodule follow-up guidelines [6], also stated that growth rate using volume was more sensitive to tumor growth than the current practice using nodal diameter [7,8,9]. However, the same guideline acknowledges the usefulness of VDT, but stated that it was not always widely used in clinical practice and would play a greater role in the future [6].

Recent developments in deep learning and research on its use in diagnostic imaging had begun to increase the number of commercially available diagnostic imaging AIs. In the current study, we used Plus.Lung.Nodule which was produced by Plusman LLC. The AI was a diagnostic imaging support program that has obtained manufacturing and marketing approval as stipulated by the Pharmaceutical Affairs Law of Japan in May 2019 and was commercially available. The results of the performance evaluation test were the highest among the results of other published papers that could be compared [10], and the clinical use of the program was progressing in Japan. Plus.Lung.Nodule processes all the voxels of the image data input, and can display regions of interest and automatically measure their diameter, area, and volume. In addition, the program’s auto-tracking function automatically links all region of interests to previously processed images to obtain time-series data. The time series data also includes the VDT. The auto-tracking function allows VDT to be measured for all nodes without explicit manipulation by the diagnostician, thus, for example, displaying nodes with VDT <400 days [4,5], which is an indication of malignancy.

We would examine whether the use of pulmonary nodule AI could accelerate the imaging diagnosis of tumors with a confirmed histopathological diagnosis of primary lung cancer. We would like to examine not only whether or not it was possible to detect the lesion earlier than the diagnostic imaging physician by simply picking up the lesion in the examination image, but also whether or not it was possible to make an earlier diagnosis than conventional imaging diagnosis even after confirming that the tumor had a VDT < 400. Furthermore, we would examine the actual patient benefit by interpolating the results of the NELSON study, which showed the prognostic benefit of early detection by lung cancer screening.

## Materials and Methods

### Data

Figure 1 showed a data flowchart of selection of the data included, and table 1 showed a demographics of the cases. Of the 2,359 patients who underwent pulmonary resection for pulmonary nodules at the Department of Pulmonary Surgery, University Hospital in Tokyo between January 1, 2015 and December 31, 2019, 91 patients received preoperative therapy, 170 patients were not pathologically diagnosed with malignancy, 66 patients had multiple neoplastic lesions, and 12 patients were excluded based on the following criteria (198 cases diagnosed with metastasis from multiple organs), 1,835 cases were extracted. Of these, 59 patients who had undergone multiple surgeries (121 surgeries, 122 tumors) were included in the analysis. Of the 122 tumors, 30 tumors were included in the scope of primary endpoint, excluding those that did not fit the analysis (Those for which it was difficult to compare with AI, e.g., those for which no images prior to the first image were available to confirm the tumor due to transfer from another hospital.).

**Table 1.** Demographics of the cases. Abbreviations: ad., adenocarcinoma; sq., squamous cell carcinoma, respectively.

|  | Scope of analysis (121cases) | Included in primary endpoints<br>(29 cases) |
| --- | --- | --- |
| Number of nodules | 122 | 30 |
| Number of Patients | 59 | 23 |
| %Male | 58% | 69% |
| Ave. Age | 72.1 | 71.9 |
| Pathological diagnosis |  |  |
| %ad. | 85.1% | 89.7% |
| %sq. | 13.2% | 10.3% |
| %other | 1.7% | 0.0% |

**Table 2:**
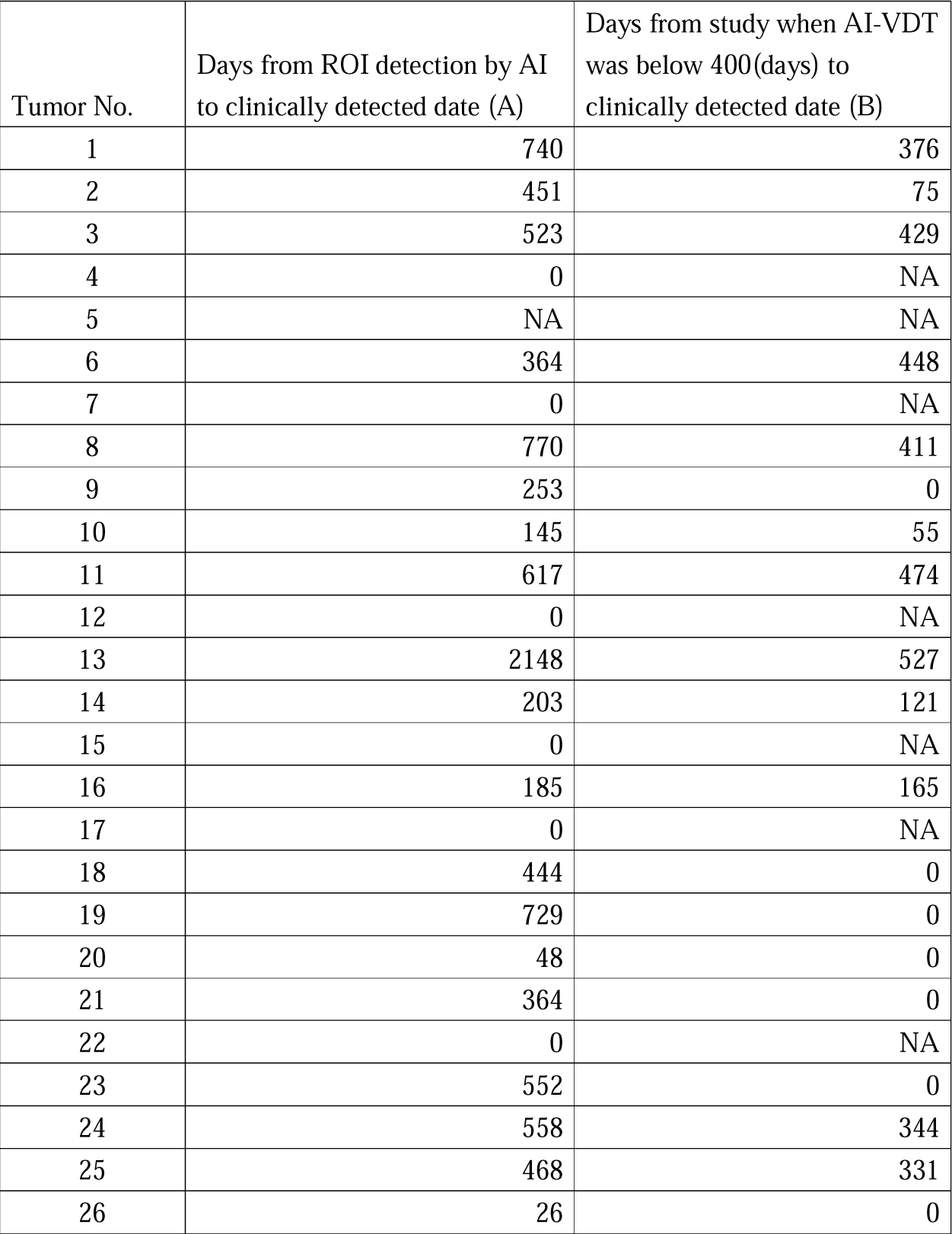
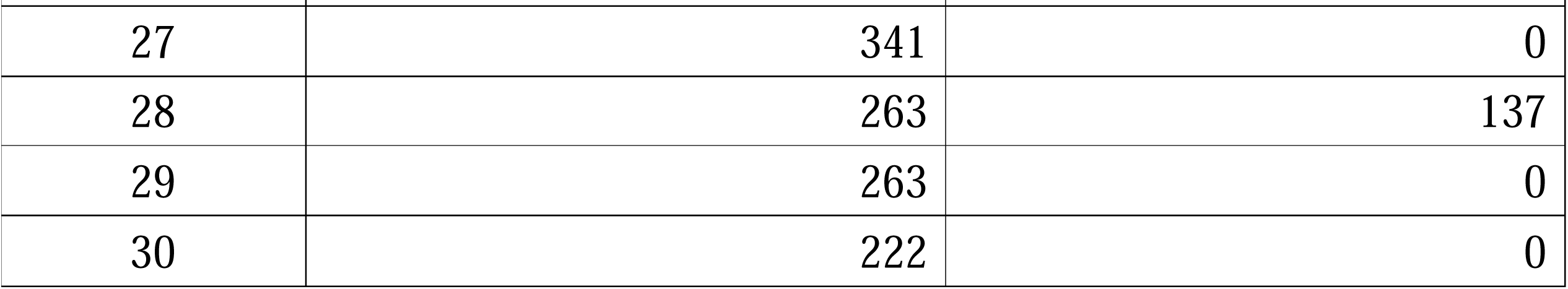
Detailed analysis results. Abbreviations: ROI, region of interest; AI, pulmonary nodule artificial intelligence; VDT, volume doubling time, respectively. column (A): NA refers to tumors for which the AI could not detect the ROI before the ROI was noted on the report. column (B): NA refers to tumors for which VDT could not be calculated before the VDT was noted on the report.

| Tumor No. | Days from ROI detection by AI to clinically detected date (A) | Days from study when AI-VDT was below 400(days) to clinically detected date (B) |
| --- | --- | --- |
| 1 | 740 | 376 |
| 2 | 451 | 75 |
| 3 | 523 | 429 |
| 4 | 0 | NA |
| 5 | NA | NA |
| 6 | 364 | 448 |
| 7 | 0 | NA |
| 8 | 770 | 411 |
| 9 | 253 | 0 |
| 10 | 145 | 55 |
| 11 | 617 | 474 |
| 12 | 0 | NA |
| 13 | 2148 | 527 |
| 14 | 203 | 121 |
| 15 | 0 | NA |
| 16 | 185 | 165 |
| 17 | 0 | NA |
| 18 | 444 | 0 |
| 19 | 729 | 0 |
| 20 | 48 | 0 |
| 21 | 364 | 0 |
| 22 | 0 | NA |
| 23 | 552 | 0 |
| 24 | 558 | 344 |
| 25 | 468 | 331 |
| 26 | 26 | 0 |

| Tumor No. | Days from ROI detection by AI<br>to clinically detected date (A) | Days from study when AI-VDT<br>was below 400(days) to<br>clinically detected date (B) |
| --- | --- | --- |
| 27 | 341 | 0 |
| 28 | 263 | 137 |
| 29 | 263 | 0 |
| 30 | 222 | 0 |

**Table 3:** Summary of statistical analysis. Abbreviations: AI, pulmonary nodule artificial intelligence; VDT, volume doubling time, respectively.

|  | Number of tumors<br>detected by AI prior to<br>radiology report | Number of tumors<br>detected by trigger of<br>AI-VTD(<400days)<br>prior to radiology report |
| --- | --- | --- |
| Number of tumors | 23 | 13 |
| proportion<br>(p-value) | 77%<br>(<0.05) | 43%<br>(<0.05) |
| Maximum days | 2148 | 527 |
| Minimum days | 26 | 55 |
| Average days | 464 | 299 |

**Figure 1.**
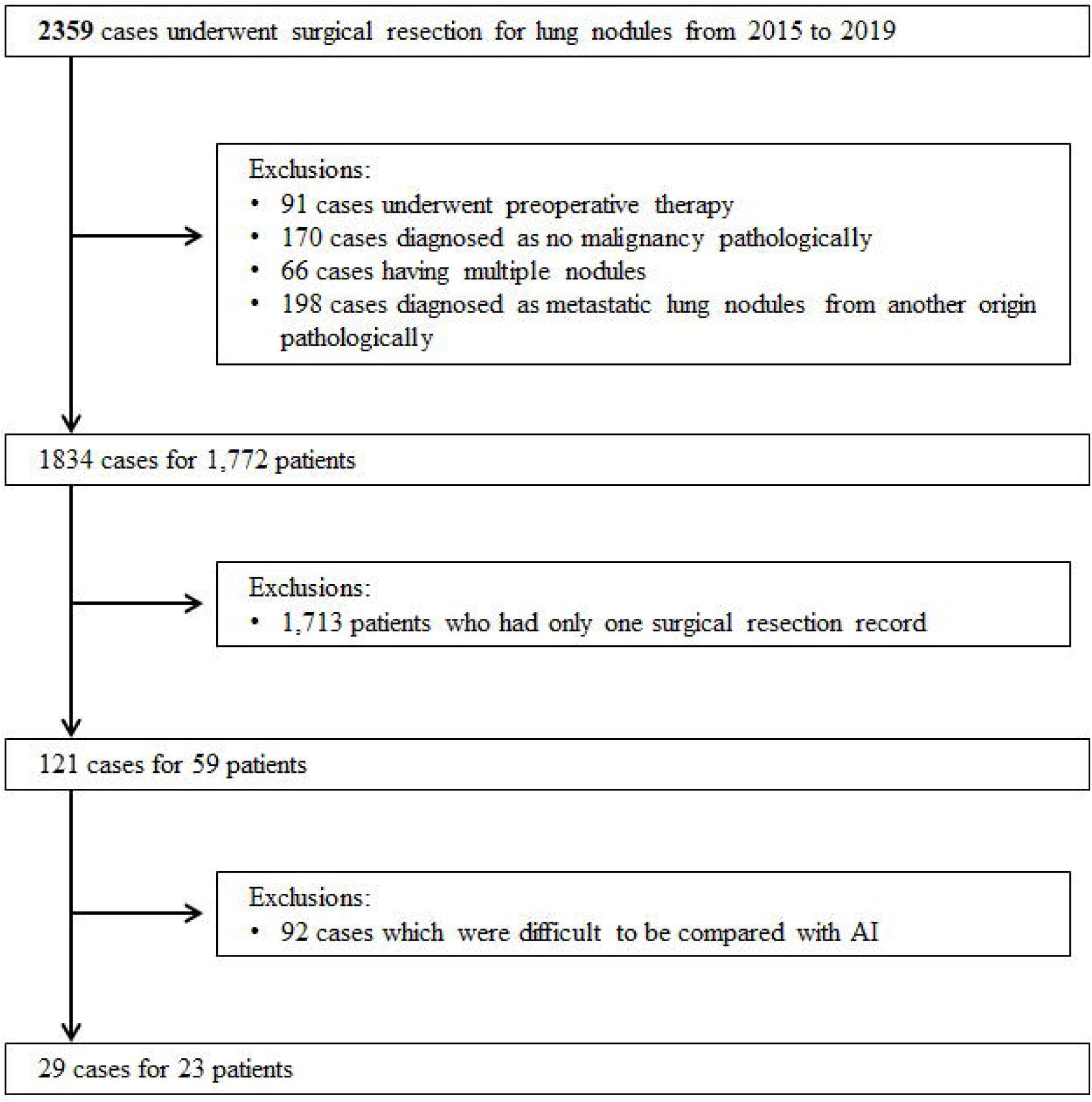
Data flowchart. Abbreviations: AI, pulmonary nodule artificial intelligence.

### Specification of target nodules

The lesion to be evaluated was identified on CT images based on the surgical record, pathology report, and diagnostic imaging report, and the date of examination of the test noted on the diagnostic imaging report was the date of discovery of the tumor in question. The description on the diagnostic imaging report was certified to the extent that it could be considered in daily practice that the diagnostic imaging physician recognized and pointed out the lesion, regardless of whether it was benign or malignant, and whether it was described in the findings or diagnosis section. In cases where multiple lesions were found within the area of interest (e.g., lower right lobe), the key image attached to the diagnostic imaging report was used to determine which lesion was the one that was noted. All imaging reports were read by radiologists, including two or more imaging specialists.

The AI, on the other hand, processed all previous images and used the auto-tracking function to detect the nodule to be analyzed and to measure the VDT (AI-VDT). Whether or not the AI found a tumor on each examination image was determined by whether or not the AI could reasonably determine which tumor was pointed to by surrounding the outline of the tumor in question and not overlapping with other lesions.

### Analysis

The date of the examination for each tumor with an AI-VDT below 400 Days was used as the AI discovery date, and was compared with the discovery date of the imaging diagnosis. The primary endpoint was the percentage of earlier detection compared to radiology report. Statistical analysis was performed with a binomial test (one-tailed) with an effect size margin of 10%.

In addition, the mortality reduction effect of early detection of lung cancer was estimated using the follow-up [3] data from the NELSON study, using the following interpolation formula.

Effectiveness of this medical device in reducing 10-year lung cancer mortality

= A. 10-year lung cancer mortality reduction in the group corresponding to stages I-IIIA in the NELSON trial (34.9%)

x (Proportion of patients with early detection / B. The same proportion in the group corresponding to stages I-IIIA in the NELSON trial (67%))

x (its average number of days of early progression / C. The same average number of days in the NELSON study (1,191Days))

## Results

### Summary of results

The results showed that in 13 of the 30 cases (proportion 43%: P value < 0.05) the tumor could be noted earlier than actual reported date. The distribution of earlyization was a minimum of 55 days, a maximum of 527 days, and a mean of 299 days.

In a subanalysis, 23 of 30 patients (77%: P value < 0.05) had tumors that could be detected earlier than by diagnostic imaging, regardless of VDT, when the date of AI detection was used as the date of detection. The distribution of earlyization was a minimum of 26 days, a maximum of 2,148 days, and a mean of 464 days.

In addition, using the follow-up results of the NELSON study [3], the estimated reduction in 10-year lung cancer mortality with this medical device was calculated to be 5.6%.

### Representative case

Figure 2 shows a representative case. The patient had a resection of the left lower lobe, and the main attention would have been on the recurrence at the margin, pleural thickening at the left lung base, and the other nodule that had appeared in the right lung S6 interlobar space. There was an exacerbation of interstitial pneumonia (three month prior to baseline date) on the dorsal side of the right lower lobe next to this nodule, which had disappeared on the baseline date and consecutive studies.

**Figure 2.**
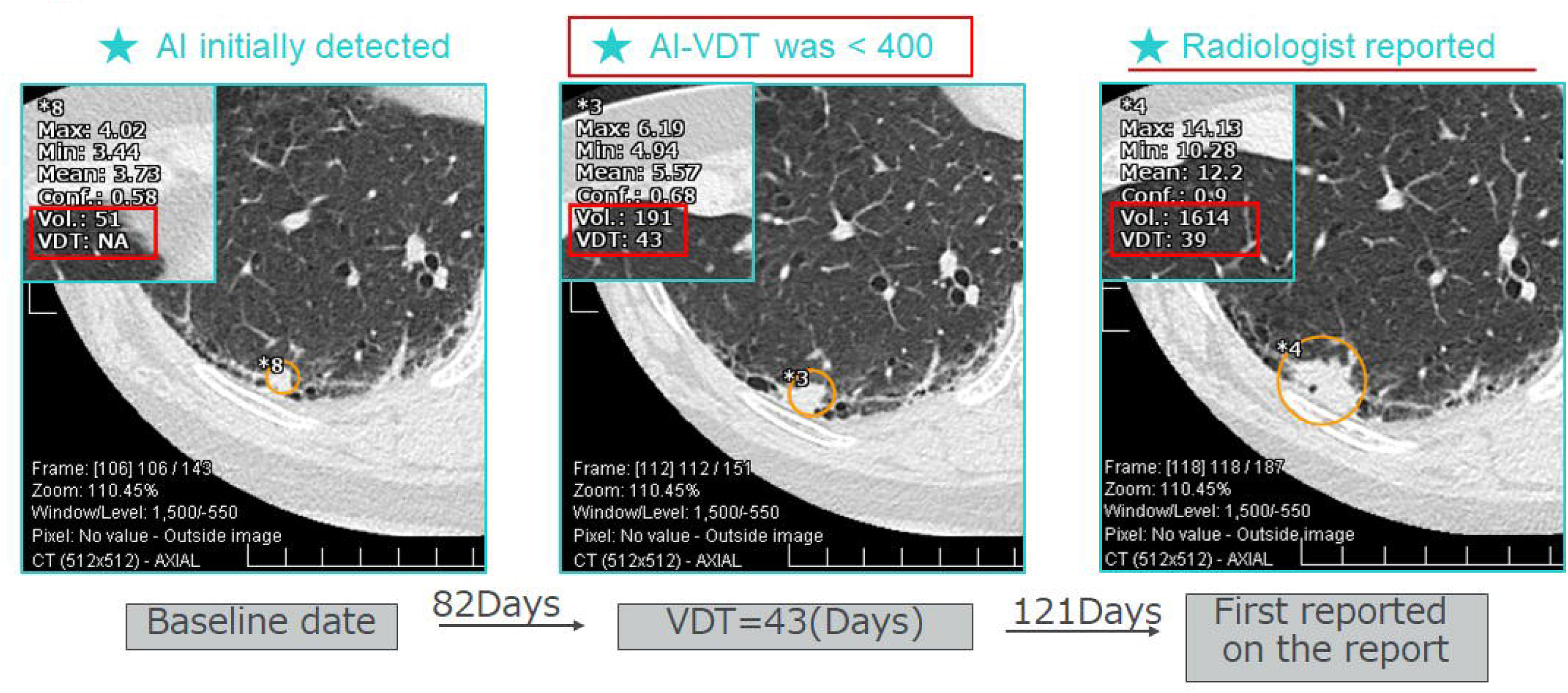
Representative case. Abbreviations: AI, pulmonary nodule artificial intelligence; VDT, volume doubling time, respectively. In this case after resection of the left lower lobe, the main attention was paid to the recurrence at the margin, pleural thickening at the left lung base, and the nodule that appeared in the right lung S6 interlobar space. There was an exacerbation of interstitial pneumonia (three month prior to baseline date) on the dorsal side of the right lower lobe next to this nodule, which had disappeared on the baseline date and consecutive studies.

A small nodule displayed in the left side of Figure 2 was overlooked in the baseline study (left). It was overlooked on the next follow-up study (middle), however at this point the AI had detected by VDT that the nodule was exacerbating at a very fast rate; if the AI had been used, it would have been possible to point it out at this point. The nodule was actually pointed out 121 days later (right). It was finally diagnosed squamous cell carcinoma.

## Discussion

### Limitation

The current study had the following limitations due to the fact that it was a study of tumors with a confirmed pathology diagnosis and that it was a retrospective real-world study.

1. The number of false positives was not included in the consideration because the pathology diagnosis for all lesions were not available for references. Such false positives include, for example, inflammatory lesions.
2. The current study was based on real-world medical data from a university hospital in Tokyo. Therefore, the competence of the radiologists who performed the readings was not controlled in the sense of conducting a study. For example, the number of years of experience as radiologists varied.
3. The current study was conducted as a retrospective study, and it was not possible to collect significant evidence on whether the intervention period would have been earlier if pulmonary nodule AI had been used in clinical practice. In this regard, based on the facts of the current study as a voluntary questionnaire to radiologists and respiratory surgeons, a probability of greater patient benefit than before was ensured if pulmonary nodule AI had been used clinically. In addition, we intend to conduct a large-scale prospective study to strengthen the mentioned qualitative evaluation in the future.

### Clinical advantages

In general, with respect to lung cancer, the probability of malignancy of tumors with VDT below 400 Days is thought to be high, and tumors with VDT above 500 Days are overwhelmingly benign nodules [4,5]. It has long been recognized that lung lesion VDT is useful in assessing malignancy. However, to measure VDT, it was necessary to measure the volume by enclosing the contour outline of the lesion in three dimensions on image data from at least two examinations at different time points. Such segmentation of a 20 mm tumor on a CT image with a slice thickness of 2 mm would require surrounding the contour outline of the tumor on approximately 10 CT slice images. Tumor volume measurement requires manual segmentation by a physician, but it was not actively implemented in clinical practice due to its inherent limitations in accuracy and reproducibility, both of which were critically important. In fact, none of the imaging reports of the 59 patients included in the current study were identified as having volume measurements.

In the current study, we confirmed that VDT was calculated for all detected lung nodule candidates using the auto-tracking function of lung nodule AI and pointed out by a threshold of 400 days, which could point out primary lung cancer earlier than conventional imaging diagnosis. In other words, they showed that the VDT, which had already been established as a follow-up indicator for nodules [6], could be “automatically applied to all potential nodules” using AI, bringing medical benefits beyond the limitations of conventional imaging diagnosis.

In conventional clinical practice, even if VDT was used for follow-up of nodules that had been selected for follow-up (which was already rare), the application of VDT to all potentially existing nodules before they were selected for follow-up would be extremely inefficient and would significantly impair the efficiency of diagnostic imaging. This would seriously impair efficiency and go beyond the scope of diagnostic imaging. The use of lung nodule AI enables follow-up of nodules that would not normally be subject to VDT calculation with VDT grading, thus increasing the chances of detecting lung cancer in stages that would otherwise go undetected by the diagnostic imaging physician.

In general, the earlier the stage, the more treatment options are available. We were able to show in the current study that the auto-tracking function of AI provides medical benefits that went beyond the limitations of conventional diagnostic imaging. It should be noted that although at least two examination images at two time points were required to calculate the VDT, even under such conditions, the detection of definite lung cancer could still be accelerated compared to conventional imaging diagnosis. Even if VDT could not be calculated at the first detection time, it might be possible to detect suspected lung cancer by the professional judgment (expert judgment) of a radiologist.

The figure 3 schematically illustrates the medical benefits of VDT measurement with the auto-tracking function of AI (AI-VDT). It shows the effect of early detection by enabling VDT measurement for all potential nodules, and shortening the follow-up period by making available the VDT grading index, which, in total, accelerates the timing of therapeutic intervention.

**Figure 3.**
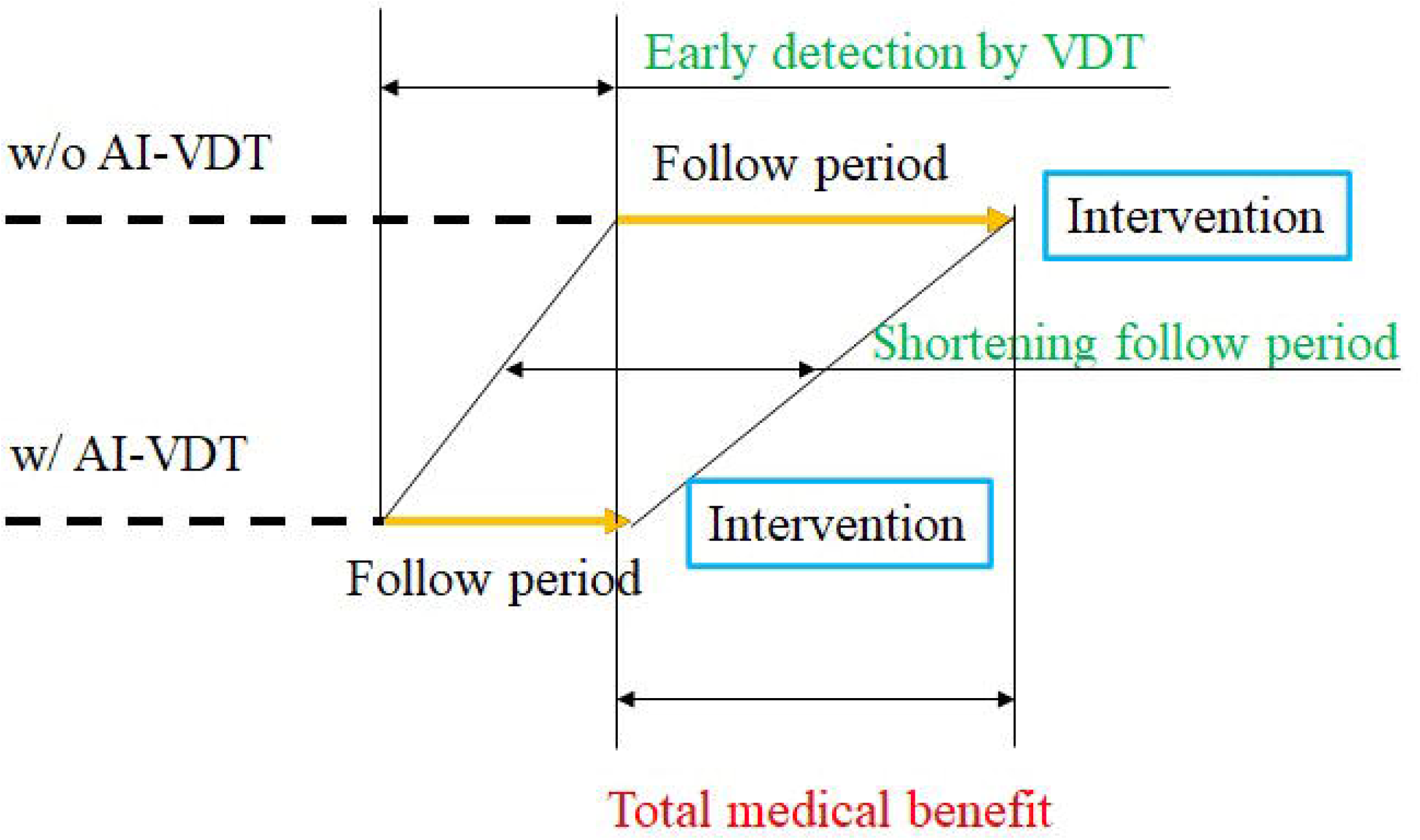
Clinical benefit schema of AI-VDT. Abbreviations: AI, pulmonary nodule artificial intelligence; VDT, volume doubling time, respectively. Schematically illustrates the clinical benefits of VDT measurement with the auto-tracking function of lung nodule AI (AI-VDT).

In, the current study, by automatically calculating the VDT of all lung lesion ROIs using the auto-tracking function of the Plus.Lung.Nodule, it could be said that (1) approximately 43% of primary lung cancers are detected an average of 299 days earlier, thereby reducing 10-year lung cancer mortality by approximately 5.6%.

## Data Availability

All data produced in the present study are available upon reasonable request to the authors

## References

1. The National Lung Screening Trial Research Team. Reduced lung-cancer mortality with low-dose computed tomographic screening. N Engl J Med 2011;365:395–409.

2. The National Lung Screening Trial Research Team. Lung cancer incidence and mortality with extended follow-up in the National Lung Screening Trial. J Thorac Oncol 2019;14:1732–1742.

3. de Koning HJ, van der Aalst CM, de Jong PA, et al. Reduced Lung-Cancer Mortality with Volume CT Screening in a Randomized Trial. NEJM; Published online 29 January 2020. DOI: 10.1056/NEJMoa1911793

4. Kanashiki Maki, Tomizawa, Takuji, Yamaguchi, Iwao, Kurishima, Koichi, Hizawa, Nobuyuki, Ishikawa, Hiroichi, Kagohashi, Katsunori Satoh, Hiroaki. Volume doubling time of lung cancers detected in a chest radiograph mass screening program: Comparison with CT screening. (2012) Oncology Letters. 4 (3): 513. doi:10.3892/ol.2012.780

5. Effect of Tumor Volume Doubling Time on Prognosis for Stage I Non–small Cell Lung Cancers No, H. et al. International Journal of Radiation Oncology • Biology • Physics, Volume 99, Issue 2, E487

6. MacMahon H, Naidich DP, Goo JM, et al. Guidelines for Management of Incidental Pulmonary Nodules Detected on CT Images: From the Fleischner Society 2017. Radiology. 2017; 284(1): 228–43.

7. Horeweg N, van der Aalst CM, Vliegenthart R et al. Volumetric computed tomography screening for lung cancer: three rounds of the NELSON trial. Eur Respir J 2013;42(6):1659–1667. Crossref, Medline, Google Scholar

8. Heuvelmans MA, Oudkerk M, de Bock GH et al. Optimisation of volume-doubling time cutoff for fast-growing lung nodules in CT lung cancer screening reduces false-positive referrals. Eur Radiol 2013;23(7):1836–1845. Crossref, Medline, Google Scholar

9. Mehta HJ, Ravenel JG, Shaftman SR et al. The utility of nodule volume in the context of malignancy prediction for small pulmonary nodules. Chest 2014;145(3):464–472. Crossref, Medline, Google Scholar

10. Kazuhiro Suzuki, Yujiro Otsuka, Yukihiro Nomura, Yukihiro Nomura, Kanako K. Kumamaru, Ryohei Kuwatsuru, Shigeki Aoki, Development and validation of a modified three-dimensional U-net deep-learning model for automated detection of lung nodules on chest CT images from the Lung Image Database Consortium and Japanese datasets. Academic Radiology DOI: 10.1016/j.acra.2020.07.030

